# Evaluating OCT Device-Reported Image Quality Score: Towards a Task-Specific Quality Gate for Deep Learning-based Outer-Retina and Choroid Boundary Segmentation

**DOI:** 10.64898/2026.05.17.26353399

**Authors:** Adarsh Gadari, Atharva Ajay Vichare, Francesca Corona, Sharat Chandra Vupparaboina, Shreyaa Rohindra Lall, Giulia Gregori, Nasiq Hasan, José-Alain Sahel, Jay Chhablani, Sandeep Chandra Bollepalli, Kiran Kumar Vupparaboina

**Affiliations:** University of Pittsburgh School of Medicine, Pittsburgh, Pennsylvania, USA; Università degli Studi di Cagliari, Cagliari, Sardinia, Italy; Polytechnic University of Marche, Ancona, Italy; Netramind Innovations Inc, USA

## Abstract

Manufacturer-defined signal-strength indices are frequently employed as quality benchmarks for automated optical coherence tomography analysis, yet their empirical relationship with deep learning segmentation accuracy remains unclear. Because these metrics were originally developed for conventional image-processing pipelines, their ability to predict modern model-based segmentation accuracy has not been empirically validated. To address this gap, we evaluated the Heidelberg Spectralis Q-score against U-Net segmentation performance across 5,047 B-scans from 103 eyes for three anatomical boundaries of the posterior segment of the eye: the Ellipsoid Zone (EZ), Bruch’s Membrane (BM), and Choroid Outer Boundary (COB). Alongside standard boundary agreement metrics (MAE, MSE, Dice Similarity Coefficient), we adapted the Earth Mover’s Distance (EMD) from optimal transport theory as a boundary evaluation metric. Unlike column-wise averages, EMD quantifies boundary agreement as a 2-D geometric displacement, directly measuring residual spatial displacement between the model segmented boundary and the ground-truth boundary. Our results demonstrate that the Q-score — originally designed to gate image-processing-based automated analysis — is a poor predictor of deep learning boundary segmentation accuracy, with explained variance (*R*^2^) failing to exceed 1.4% across all three boundaries. We further observed a monotonically increasing error hierarchy with anatomical depth (EZ < BM < COB), consistent across metrics, which is unexplained by the signal strength. At the COB, correlations were paradoxically positive, explained by a B-scan-level mediation chain: higher Q-scores correspond to greater choroidal thickness (*r* = 0.113, *ρ* = 0.158), which in turn predicts higher COB segmentation error (*r* = 0.165, *ρ* = 0.191) — a localization difficulty that global signal strength cannot capture. Collectively, these findings challenge the implicit assumption that signal-strength-based quality thresholds are a reliable proxy for deep learning model performance, and motivate a shift toward task-specific acquisition quality criteria calibrated to model performance rather than signal interpretability.

## 1. Introduction

Optical Coherence Tomography (OCT) has become the standard imaging modality for non-invasive, high-resolution visualization of retinal microstructure in clinical ophthalmology [1, 2]. Among the anatomical boundaries of clinical interest, the Ellipsoid Zone (EZ), Bruch’s Membrane (BM), and Choroid Outer Boundary (COB) have emerged as critical biomarkers for conditions such as Age-related Macular Degeneration (AMD), Central serous chorioretinopathy (CSCR), and macular telangiectasia (MacTel) where their precise quantification directly informs disease staging, treatment monitoring, and progression assessment [3, 4]. Figure 1 (left) shows the different segments of the OCT scan: Vitreous Chamber, Retina, Choroid and Sclera. Figure 1 (right) shows all three boundaries on a representative B-scan: EZ at the inner retina, BM at the retina–choroid junction, and COB at the choroid–sclera interface, ordered from superficial to deep.

**Fig. 1.**
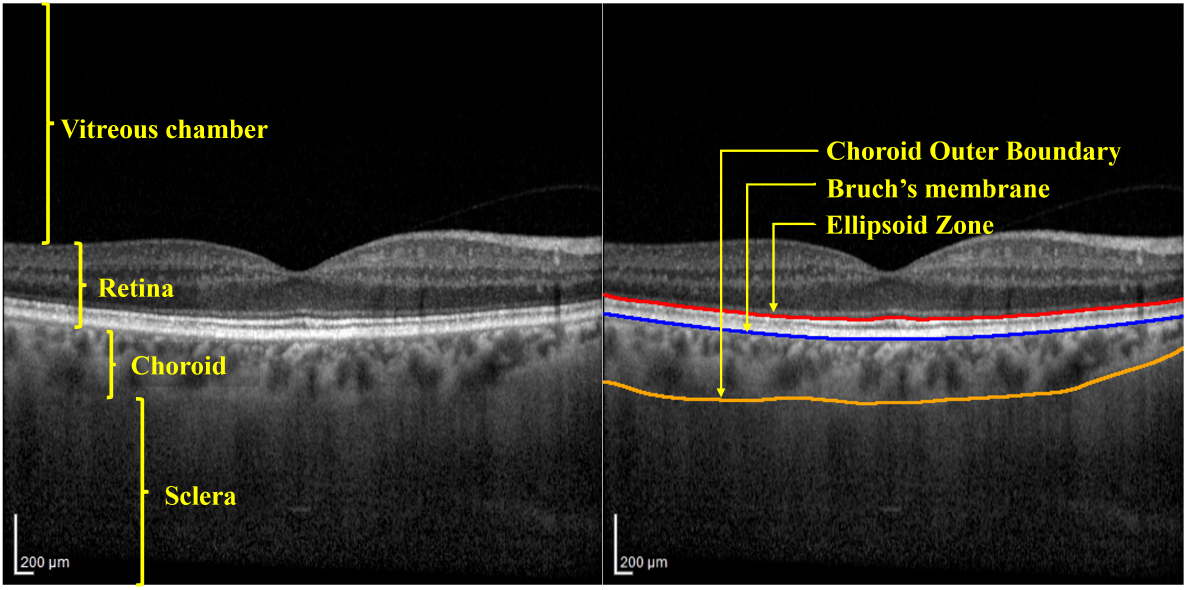
Cross-sectional OCT B-scan of the posterior retina, oriented with the inner retina at top and the choroid below. Brighter regions reflect stronger signal return. Three boundaries are marked in order of anatomical depth: the Ellipsoid Zone (EZ, red) — a bright hyperreflective band at the photoreceptor layer; Bruch’s Membrane (BM, blue) — the thin basement membrane separating the retina from the choroidal vasculature; and the Choroid Outer Boundary (COB, orange) — the outer limit of the choroid at the choroid–sclera interface, where signal progressively attenuates with depth.

Manual segmentation of these boundaries across OCT volumes is time-intensive and subject to inter-grader variability, creating significant bottlenecks in both clinical workflows and large-scale research studies. Deep learning (DL) architectures — particularly convolutional neural networks (CNNs) — have demonstrated segmentation accuracy comparable to expert graders while substantially reducing analysis time [5–7]. Figure 2 shows a representative example: model predictions and expert ground truth overlaid on a single B-scan for all three boundaries. Despite these advances, deploying DL models in clinical practice raises a practical but underexplored question: how should clinicians decide whether a given scan is of sufficient quality for automated segmentation to be trusted?

**Fig. 2.**
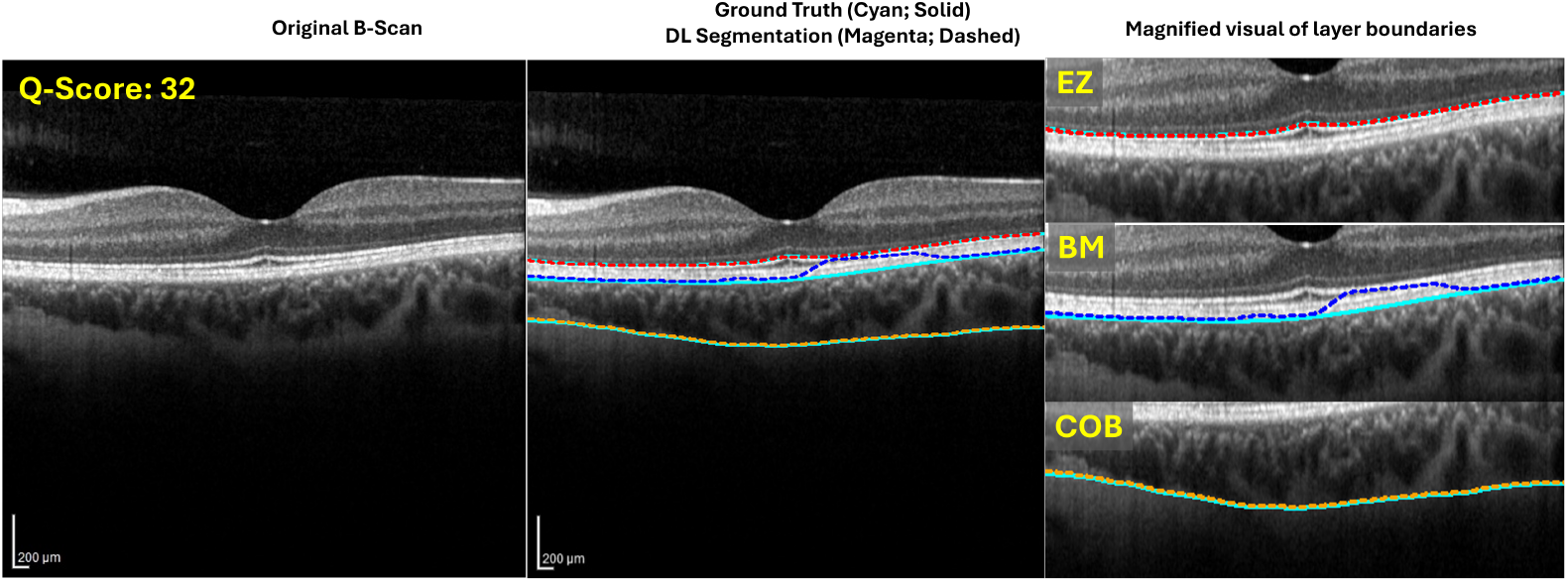
Deep learning boundary segmentation versus expert ground truth on a representative B-scan (Q-score = 32, well above the clinical inclusion threshold of ≥ 15). *Left:* original B-scan. *Center:* expert ground truth (cyan, solid) and model predictions (dashed) overlaid — EZ in red, BM in blue, COB in orange. *Right:* magnified view showing regions of close agreement and boundary-level divergence between ground truth and model output. Solid cyan and dashed colored lines denote ground truth and model predictions respectively in all subsequent comparison figures.

Current clinical practice addresses acquisition quality through device-generated signal-strength indices, a convention common across OCT platforms. The Heidelberg Spectralis — one of the most widely deployed platforms in clinical and research settings, and the prototype for this study — reports one such index per B-scan: the Heidelberg Quality score (Q-score). While a Q-score of ≥ 15 is the widely adopted threshold for traditional automated analysis [8], stricter thresholds of ≥ 20 [9, 10] or ≥ 25 [11] are frequently employed as inclusion criteria in clinical research to ensure high data integrity. Traditional automated analysis relies on global image properties — signal strength, contrast, and brightness — that Q-score directly reflects. Deep learning models, however, operate differently: a CNN trained to identify retinal boundaries extracts hierarchical spatial features through learned convolutional filters, relying on local contrast, edge definition, and spatial relationships at specific anatomical interfaces. The urgency of resolving this mismatch has sharpened with the entry of OCT-derived boundary metrics into regulatory endpoints: the FDA approval of revakinagene taroretcel (Encelto) for macular telangiectasia type 2 was supported by Phase 3 trials in which rate of change in EZ area loss was the primary outcome measure [12]. When automated boundary segmentation feeds directly into a trial endpoint, an image that passes a signal-strength threshold but yields unreliable segmentation introduces silent measurement error into the evidence base on which treatment approvals depend.

Efforts to move beyond manufacturer-defined thresholds toward standardized quality control have been most systematically developed for retinal OCT. The OSCAR-IB consensus criteria defined seven human-led quality dimensions: obvious errors, signal strength, centration, algorithm failure, retinal pathology, illumination, and beam placement [13]. Because human-calibrated criteria are grounded in the same global image properties and perceptive image quality that DL models do not primarily rely on, the OSCAR IB criteria do not transfer directly to AI pipelines. This gap is addressed by proposing the OSCAR-AI framework [14], which calls for task-appropriate quality standards in algorithmic workflows. However, neither framework empirically characterizes whether acquisition metrics predict model performance on specific segmentation tasks. A parallel line of work has instead pursued automation itself — replacing human graders with learned quality models. For instance, AQUA-OCT proposed a modular neural network system to automate and standardize OCT quality assessment, targeting replacement of manual OSCAR-IB rating across scan centering, signal quality, and image completeness [6]. While the system includes heuristic checks for surface segmentation plausibility, it does not optimize quality criteria as predictors of downstream segmentation performance. Most recently, ROQUS introduced a ranking-based CNN producing a continuous quality score from pairwise B-scan comparisons [15]; while it represents the current state of the art — achieving 0.85 ROC-AUC in flagging acquisition issues and approaching inter-human agreement on quality rankings — its usability criterion is operationalized as human-judged layer readability and diagnosability. The authors identify the prediction of downstream automated analysis failure as an unvalidated open direction [15].

Across this body of work, a consistent pattern emerges: quality metrics — whether manufacturer-defined scalar thresholds, consensus human criteria, automated neural classifiers, or ranking-based learned metrics — are defined and validated against human-interpretable image properties or image-level classification tasks. Whether acquisition quality indicators predict the performance of deep learning models on a precise retinal boundary segmentation task remains, to our knowledge, unexamined. We address this gap using the Heidelberg Q-score as a prototype for signal-strength-based quality gating in clinical AI pipelines.

In this paper, we characterize this relationship through a retrospective study applying two validated U-Net-based models to segment the EZ, BM, and COB across 5,047 B-scans from 103 healthy eyes. Segmentation accuracy was evaluated against device-reported Q-scores using a multi-metric framework; alongside standard boundary agreement metrics, we introduce the Earth Mover’s Distance (EMD) from optimal transport theory as a geometric boundary evaluation measure that captures total spatial displacement rather than column-wise averaging.

This study yields five contributions spanning evaluation methodology and clinical governance of AI-assisted OCT analysis:

- **An empirical characterization of Q-score as a quality gate for DL segmentation**. We demonstrate that device-reported signal strength is not predictive of deep learning boundary segmentation accuracy across any of the three anatomical boundaries examined and across any metric family.
- **A novel adaptation of Earth Mover’s Distance for OCT boundary segmentation evaluation**. EMD, established in computer vision for shape matching, is adapted here to quantify total geometric boundary displacement between predicted and ground-truth contours — capturing spatially coherent shifts that column-wise metrics such as MAE cannot represent.
- **Identification of anatomical depth as the primary driver of segmentation error**. Segmentation error increases monotonically from EZ to BM to COB in a pattern consistent across independent metric families, reflecting boundary-intrinsic localizability rather than image signal quality.
- **A mechanistic explanation for the paradoxical COB finding**. At the deepest boundary, higher Q-scores correspond to greater choroidal thickness, which in turn predicts harder COB localization — a failure mode that global signal strength cannot capture.
- **A clinical governance argument for task-specific quality criteria**. The manufacturer-defined Q-score threshold widely used to gate automated OCT analysis was designed for traditional image-processing pipelines and has never been validated against DL model performance — constituting a silent failure risk in clinical deployment.

These findings motivate a fundamental shift from signal-strength-based quality gates toward task-specific acquisition quality criteria calibrated directly to segmentation model performance.

## 2. Methods

### 2.1. Dataset

This retrospective study was conducted at the University of Pittsburgh Medical Center, USA, with approval from the Institutional Review Board of the University of Pittsburgh Medical School. All procedures adhered to the tenets of the Declaration of Helsinki. Subjects underwent OCT examination of the posterior pole on a Heidelberg Engineering Spectralis system (Heidelberg Engineering, Heidelberg, Germany) following the Enhanced Depth Imaging (EDI) protocol [16]. Each volume covered a scan area of 6 × 6 mm^2^ and a depth of 2 mm, yielding 49 B-scans per volume at an image resolution of 496 × 512 pixels. The dataset comprised 103 healthy eyes from 67 subjects (male: 20, female: 47; age range: 18–80 years; age: 45 ± 17.8 years), resulting in 5,047 B-scans in total. Subsequently, device-reported Q-scores for each B-scan were extracted from the accompanying DICOM and XML files exported via Heidelberg’s Heyex Explorer 2 PACS software [17]. Across the dataset, Q-scores had a mean of 27.65 (± 3.86), spanning a range of 13–43.

### 2.2. Boundary Segmentation

#### 2.2.1. Deep Learning Models

Automated segmentation of EZ and BM was performed using the NetraMind Innovations Outer Retinal Analyzer (NMI-ORA^®^) [18], a suite of independent U-Net-based boundary segmentation models [19], each trained and validated for a single boundary. The COB was segmented using a U-Net-based Choroid Segmentation model (NMI-ChoroidAI^®^) [20]. Each model receives a B-scan as an input and returns a binary boundary mask. To ensure spatial continuity, extracted boundaries were smoothed using Locally Weighted Scatterplot Smoothing (LOESS) [21] with a window of 0.1, yielding a continuous per-boundary prediction across the full width of each B-scan. Figure 2 shows a representative B-scan with ground truth (cyan, solid) and model predictions (dashed) overlaid for all three boundaries; this color convention is maintained in all subsequent comparison figures.

### 2.3. Evaluation Framework

To comprehensively assess the relationship between image quality and segmentation performance, each B-scan was evaluated using metrics spanning three complementary mathematical perspectives.

#### 2.3.1. Moment-Based and Order Statistics

Let *ŷ*_*i*_ and *y*_*i*_ denote the predicted and ground-truth boundary pixel locations at the *i*-th A-scan. The absolute error per A-scan is |*e*_*i*_ | = | *y*_*i*_ − *ŷ*_*i*_ |.

Mean Absolute Error (MAE) per B-scan over *n* A-scans:

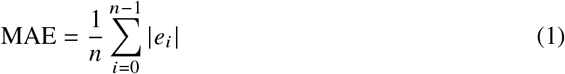

Standard Deviation of Absolute Error (SDAE):

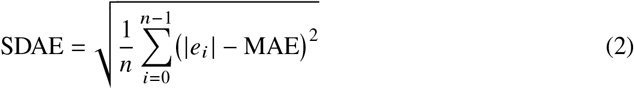

Coefficient of Variation (CV) [22, 23]:

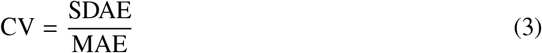

Order statistics per B-scan:

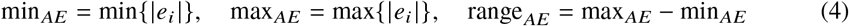

#### 2.3.2. Dice Similarity Coefficient

To evaluate boundary agreement as a regional overlap while accounting for minor axial displacements, we utilize a Dilated Dice Similarity Coefficient (DSC_*δ*_) [24]. Unlike standard area-based Dice, this approach restricts the evaluation to a specific tolerance zone—or “ribbon”—surrounding the boundary.

Each predicted boundary 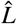 and ground-truth boundary *L* is first transformed into a binary mask. For a given dilation margin *δ*, a mask *M*_*δ*_ is defined as the set of all pixels *p* whose Euclidean distance to the boundary is less than or equal to *δ*:

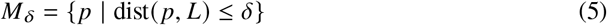

Let *A*_*δ*_ and *B* _*δ*_ represent the binary masks generated by dilating the predicted and ground-truth boundaries, respectively, by a margin *δ* ∈ {3, 5} pixels. The Dilated DSC is then computed as:

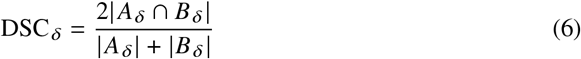

where | · | denotes the count of foreground pixels (the area of the dilated ribbon).

By varying *δ*, this metric provides a tunable measure of “forgiveness” for spatial jitter. While the general DSC formula remains the standard for measuring overlap, the dilation operation ensures the metric is specifically sensitive to the topological alignment of the thin layers characteristic of OCT imaging, rather than being dominated by the large background or foreground areas typical of traditional volumetric segmentation.

#### 2.3.3. Transport Metric: Earth Mover’s Distance

To quantify the total geometric effort required to align a predicted boundary with its ground-truth counterpart, we adopt the Earth Mover’s Distance (EMD) [25], formally equivalent to the Wasserstein-1 distance from optimal transport theory [26].

Each boundary is first extracted as a contour from its binary segmentation mask and uniformly resampled to *n* points. The predicted and ground-truth point sets, 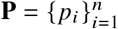 and 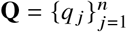, are treated as uniform discrete distributions, each assigning mass 1 / *n* to every point. To measure the cost of moving mass between any two points, we construct a cost matrix **C** ∈ ℝ^*n*×*n*^, where each entry *C*_*i j*_ = ∥*p*_*i*_ − *q*_*j*_∥_2_ is the Euclidean distance between point *p*_*i*_ in the predicted set and point *q* _*j*_ in the ground-truth set. The EMD is then defined as the minimum-cost transport plan over all valid assignments ***γ***:

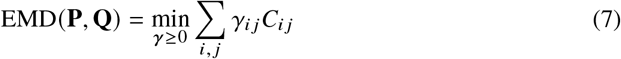

subject to ∑_*j*_ *γ*_*i j*_ = ∑_*i*_ *γ*_*i j*_ = 1 / *n*, where *γ*_*i j*_ denotes the fraction of mass transported from *p*_*i*_ to *q* _*j*_ . The optimal objective value of this minimization — the minimum total transport cost achieved at the best assignment — is a single scalar expressed in pixel units, representing the minimum total geometric displacement required to morph the predicted boundary into the ground truth.

The use of EMD as a contour similarity measure is well-established in computer vision, where it has been applied to shape matching and object recognition by treating boundaries as point distributions and measuring the minimum transport cost between them [27, 28]. We adapt this formulation to segmentation evaluation, where the transport cost between predicted and ground-truth boundary distributions directly quantifies the geometric displacement, a quantity that column-wise metrics such as MAE cannot capture because they assume fixed positional correspondence rather than finding the globally optimal match. It is therefore sensitive to spatially coherent shifts — such as a segment of the boundary being displaced laterally as a unit — that column-wise averaging may dilute by treating each A-scan error independently.

### 2.4. Statistical Analysis

Pearson correlation coefficients (*r*) were computed between per-scan Q-scores and each of the aforementioned metrics, separately for the EZ, BM, and COB boundaries. To further examine the anatomical basis of the COB findings, both Pearson and Spearman correlations were computed between per-B-scan average choroidal thickness and device-reported Q-score and COB Earth Mover’s Distance.

Given the large sample size (*n* = 5,047 B-scans), effect sizes were interpreted according to standard conventions: negligible (|*r*| < 0.1), weak (0.1 ≤ |*r*| < 0.3), moderate (0.3 ≤ |*r*| < 0.5), and strong (|*r*| ≥ 0.5) [29, 30]. It should be noted that, at this sample size, even trivially small correlations can attain statistical significance; accordingly, *p*-values are reported for completeness but effect size is treated as the primary criterion for interpreting the practical relevance of each association.

## 3. Results

### 3.1. EZ Boundary

The EZ is a bright hyperreflective band with the highest local contrast of the three boundaries, making it the most consistently localized. The NMI-ORA model achieved a mean DSC_*δ*=3_ of 0.894 and a median of 0.912 for the EZ boundary. Correlations between Q-score and segmentation performance were uniformly negligible (Table 1; Figure 3, first column). Across moment-based measures (MAE, MSE, RMSE, SDAE) and order-statistical measures (min_*AE*_, max_*AE*_, range_*AE*_), correlation coefficients fell within − 0.077 ≤ *r* ≤ − 0.015, collectively explaining under 0.6% of error variance (*R*^2^ < 0.006). CV and DSC showed weak positive associations (*r* = 0.083 and *r* = 0.095, respectively); Earth Mover’s Distance (EMD), which captures total geometric displacement rather than per-column error, confirmed the same pattern (*r* = − 0.060). Collectively, statistical significance here reflects sample size, not predictive capacity. A Q-score accounting for less than 1% of segmentation variability offers no actionable quality gate for clinical deployment.

**Table 1.** Pearson correlation coefficients (*r*) between device-reported Q-scores and per-boundary segmentation error metrics. Boldface indicates *p* > 0.05 (not statistically significant); all other entries are significant at *p* < 0.05. Statistical significance at this sample size (*n* = 5,047) reflects sample size rather than effect magnitude; effect sizes are reported in the text.

| Metric | EZ | BM | COB |
| --- | --- | --- | --- |
| MAE | −0.065 | 0.060 | 0.101 |
| MSE | −0.047 | **−0.007** | 0.063 |
| RMSE | −0.064 | 0.035 | 0.101 |
| SDAE | −0.055 | **−0.018** | 0.087 |
| MIN | −0.015 | 0.040 | **−0.001** |
| MAX | −0.077 | −0.068 | 0.096 |
| RANGE | −0.056 | −0.066 | 0.096 |
| CV | 0.083 | −0.071 | **−0.021** |
| DSC<sub>$\delta=3$</sub> | 0.095 | −0.089 | −0.087 |
| DSC<sub>$\delta=5$</sub> | 0.095 | −0.094 | −0.117 |
| EMD | −0.060 | **0.002** | 0.100 |

**Fig. 3.**
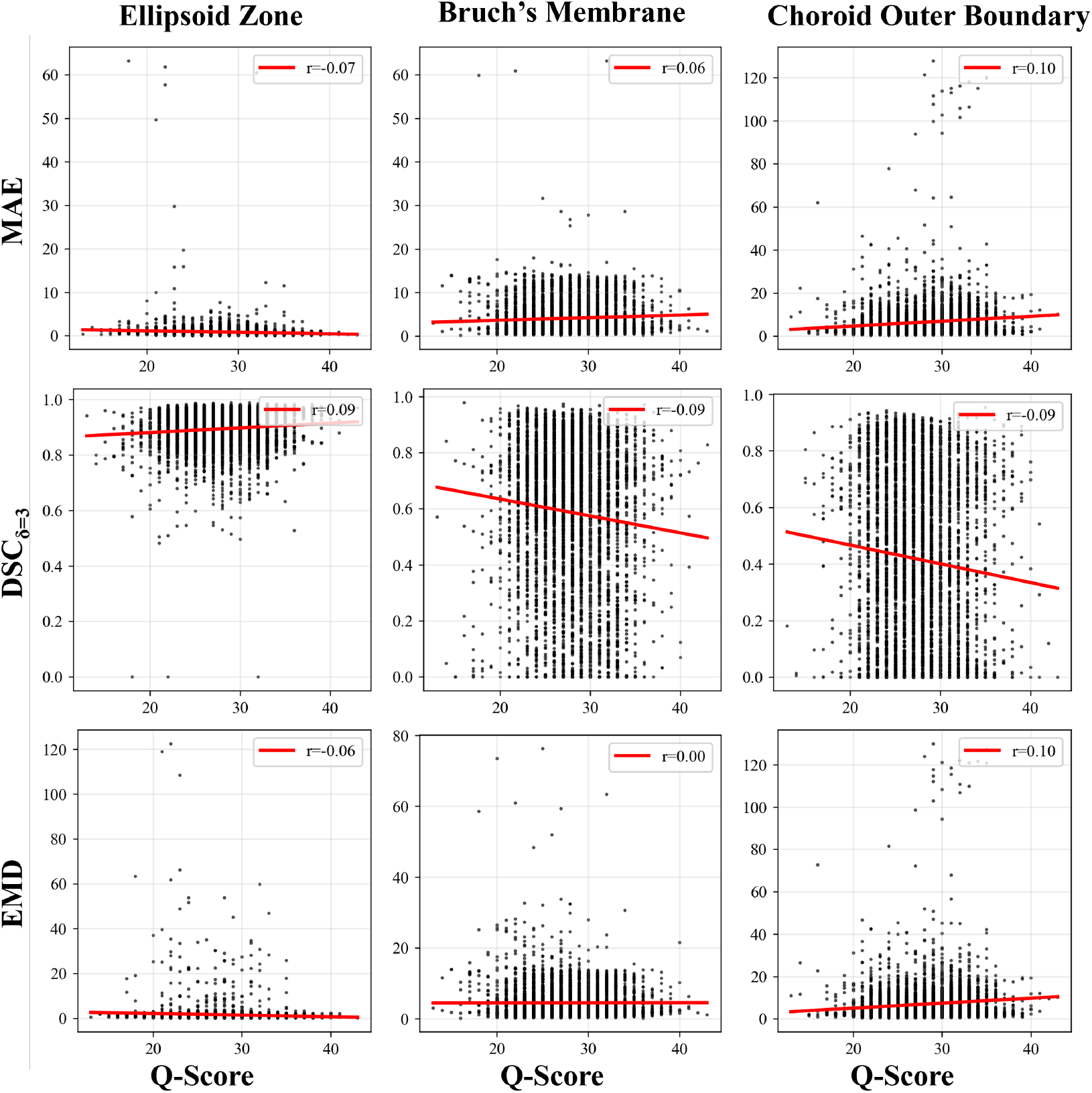
Q-score versus segmentation error across three metric families and all three anatomical boundaries (N = 5,047 B-scans per boundary). Rows represent metric families ordered from column-wise (MAE), through overlap-based (DSC_*δ*_), to geometric transport (EMD). Columns follow anatomical depth: EZ (superficial), BM (intermediate), COB (deepest). All nine panels show a flat, diffuse distribution with no discernible trend, confirming that Q-score carries no predictive information for segmentation error regardless of metric or boundary. Individual subfigure correlations are reported in Table 1.

### 3.2. BM Boundary

Unlike the EZ, the BM has no dominant reflectance peak and is localized from contrast with adjacent tissue. The BM boundary presented a more demanding segmentation target, reflected in lower performance (mean DSC_*δ*=3_ = 0.588, median = 0.658). Q-score associations nonetheless remained negligible in magnitude across all metrics (Table 1). Specifically, directional consistency was not preserved: MAE shifted from *r* = − 0.065 to *r* = + 0.060, RMSE from *r* = − 0.064 to *r* = +0.035, CV from *r* = +0.083 to *r* = −0.071, and DSC_*δ*=3_ from *r* = +0.095 to *r* = −0.089. SDAE and MSE yielded correlations indistinguishable from zero. The boundary-level EMD confirmed the pattern most directly: *r* = 0.002 (*p* = 0.87), indicating that total geometric displacement of BM contours bears no relationship to Q-score. Together, sign reversals of this kind are expected at magnitudes governed by sampling variation rather than underlying signal. They corroborate rather than contradict the EZ finding: Q-score carries no consistent directional information across boundaries.

### 3.3. COB Boundary

The choroid outer boundary is widely regarded as the most challenging to delineate due to gradual signal attenuation at the choroid–sclera interface. It yielded the lowest segmentation performance (mean DSC_*δ*=3_ = 0.416, median = 0.417) and the strongest Q-score associations among the three boundaries (Table 1). DSC_*δ*=5_ achieved a correlation of *r* = − 0.117 while MAE and RMSE each reached *r* = 0.101, the highest coefficients in this study: higher Q-scores were associated with larger segmentation errors. The minimum absolute error (min_*AE*_) yielded a near-zero association (*r* = − 0.001, *p* > 0.05). Furthermore, the boundary-level EMD reinforced the positive pattern (*r* = 0.100, *p* < 0.05). Across all metrics, *R*^2^ remained below 0.014 — the strongest associations observed in this study still explained ≈1% of segmentation variability.

### 3.4. Cross-Boundary Pattern

Across all metrics, segmentation error increased monotonically with anatomical depth. MAE followed the hierarchy EZ (0.87 px), BM (4.07 px), COB (6.39 px), consistent across all moment-based measures. EMD, a geometric displacement measure, ranked the boundaries identically: EZ (1.55 px), BM (4.51 px), COB (6.84 px). Q-score correlations showed no corresponding directional gradient, remaining within |*r*| ≤ 0.117 across all boundaries and metric families. The agreement (Figure 3) between the metrics confirms the hierarchy reflects the anatomy, not the choice of error measure.

## 4. Discussion

The central finding is unambiguous: device-reported Q-scores bear no meaningful relationship to the segmentation performance of validated deep learning models across any of the three anatomical boundaries examined. Correlations remained within |*r*| ≤ 0.117 across all metric families, with a maximum explained variance of *R*^2^ = 0.014 at the most challenging boundary. That most associations reached *p* < 0.05 is a well-characterized consequence of large-*n* inference (*n* = 5,047 B-scans) rather than evidence of a meaningful effect [29, 30]. Three lines of evidence from the boundary-level analyses reinforce this conclusion: the directional instability of associations across anatomically adjacent boundaries, the paradoxical positive direction of correlations at the COB, and the monotonic increase in segmentation error with anatomical depth in the complete absence of a corresponding increase in Q-score correlation. Figures 4 and 5 show this dissociation at the scan level: at both low and high Q-scores, accurate and inaccurate EZ and BM segmentations appear side by side.

**Fig. 4.**
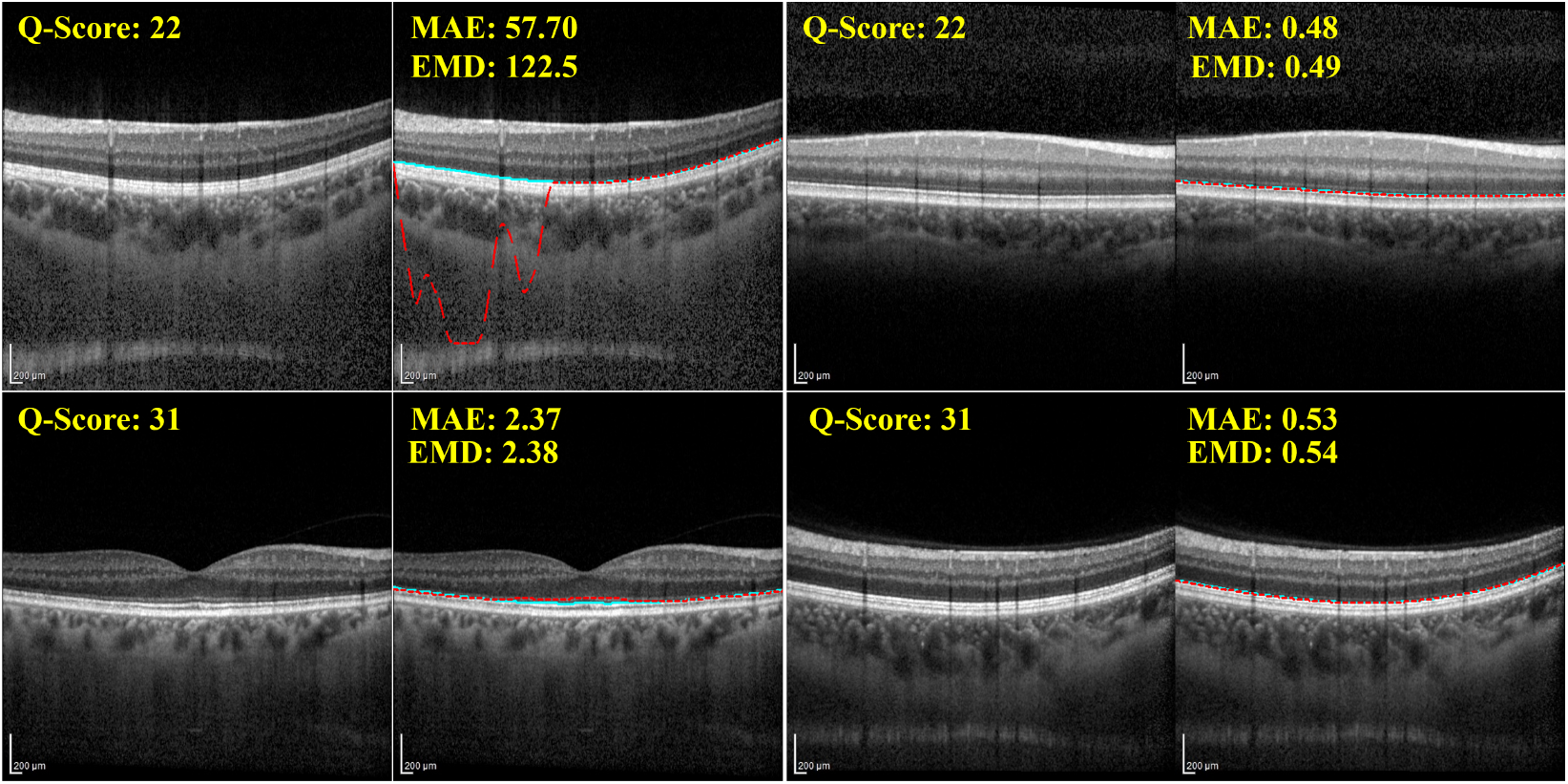
Q-score does not predict EZ segmentation outcome. Each row presents one successful and one failed segmentation at the same Q-score level: low (Q-score = 22, top row) and high (Q-score = 31, bottom row). Within each example, the left panel shows the original B-scan and the right panel shows the segmentation overlay. Ground truth: solid cyan. Model prediction: dashed red. MAE and EMD values quantify mean absolute and geometric boundary displacement respectively, per overlay. Accurate and inaccurate outcomes occur at both Q-score levels, confirming Q-score carries no predictive information for EZ segmentation accuracy.

**Fig. 5.**
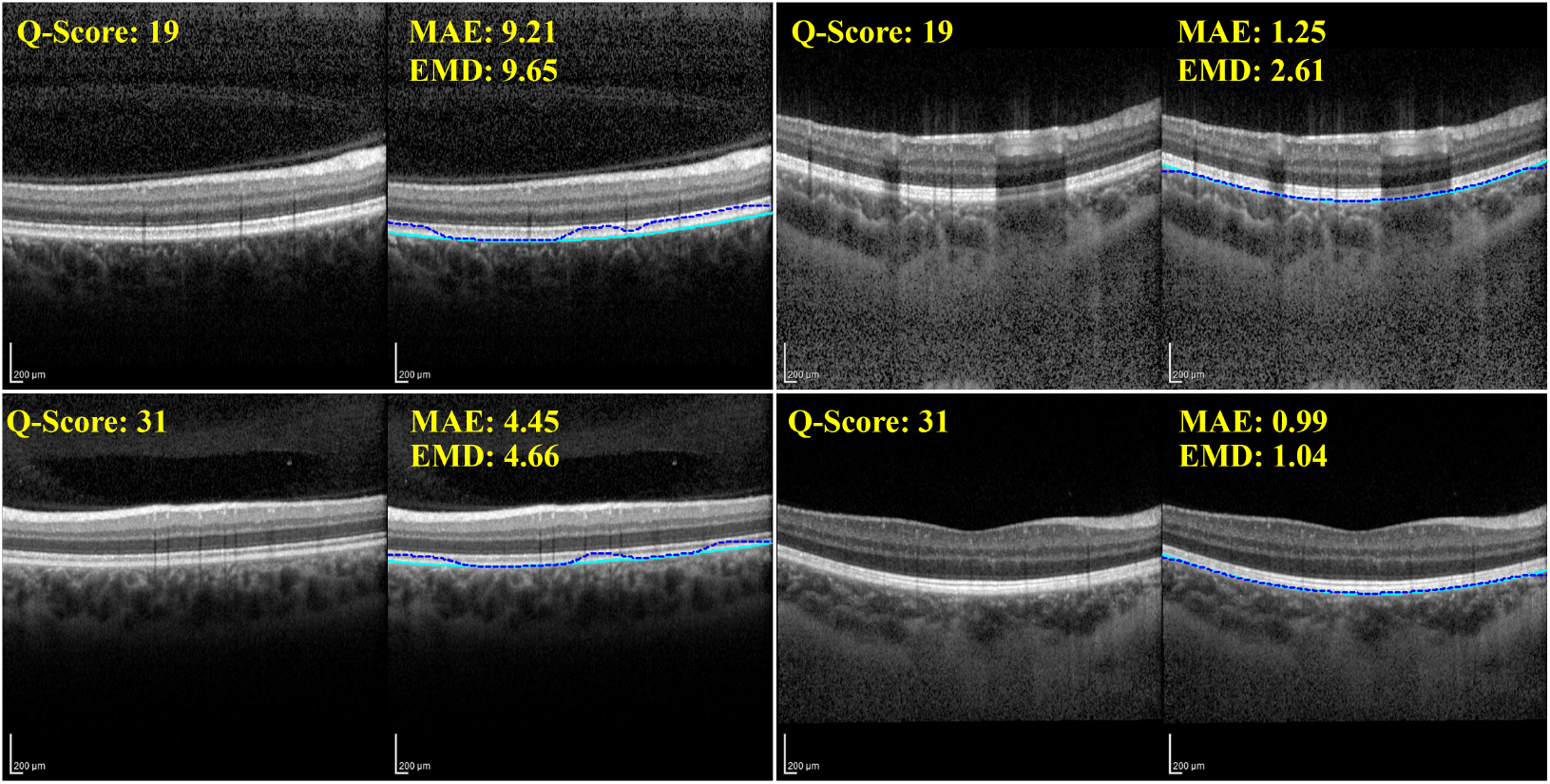
Q-score does not predict BM segmentation outcome. Each row presents one successful and one failed segmentation at the same Q-score level: low (Q-score = 18, top row) and high (Q-score = 33, bottom row). Within each example, the left panel shows the original B-scan and the right panel shows the segmentation overlay. Ground truth: solid cyan. Model prediction: dashed blue. EMD values quantify geometric boundary displacement per overlay. The sign reversal observed in Table 1 for BM — opposite in direction to EZ despite sharing the same tissue context — is consistent with Q-score carrying no directional information at either boundary.

The sign reversals across adjacent boundaries confirm that Q-score carries no consistent directional information about model error (Figures 4 and 5). For the EZ, MAE correlated negatively with Q-score (*r* = −0.065); for the BM — immediately adjacent and segmented by the same model architecture — the sign reversed (*r* = +0.060). CV and DSC_*δ*=5_ showed the same pattern: EZ (*r* = 0.083, *r* = 0.095) versus BM (*r* = −0.071, *r* = −0.094). A reliable quality indicator must consistently predict error direction across boundaries sharing the same tissue context. However, Q-score does not. The COB boundary makes a sharper case: the correlation does not merely fluctuate — it inverts.

At the COB, Q-score did not merely fail to predict segmentation error — it predicted the opposite (Figure 6). The boundary where signal attenuation is greatest was also the boundary where higher device-reported quality corresponded to larger model error. The explanation lies in the anatomy. The Q-score measures global signal strength during acquisition — but in the posterior eye, signal strength partly reflects how much tissue the beam must penetrate. The EDI protocol improves choroidal visualization by placing the choroid closer to the zero delay line, increasing signal depth [16]; in eyes where that deeper tissue is more voluminous, the returning signal may appear stronger even as the choroid–sclera interface becomes more diffuse, presenting no sharp intensity transition for the model to anchor to. This anatomical reasoning makes a testable prediction: higher Q-scores should correspond to greater choroidal thickness. The data confirm it (*r* = 0.113, *ρ* = 0.158), consistently across both Pearson and Spearman analyses. A second prediction follows directly: thicker choroids should be harder to segment at the COB. Again, the data confirm it (*r* = 0.165, *ρ* = 0.191). Each link is weak in magnitude — expected for a single factor in a multifactorial problem — but both point in the same direction. The chain is therefore: higher Q-score → thicker choroid → harder COB localization. Both better signal and the models’ difficulty are true simultaneously — and no scalar quality gate can distinguish them.

**Fig. 6.**
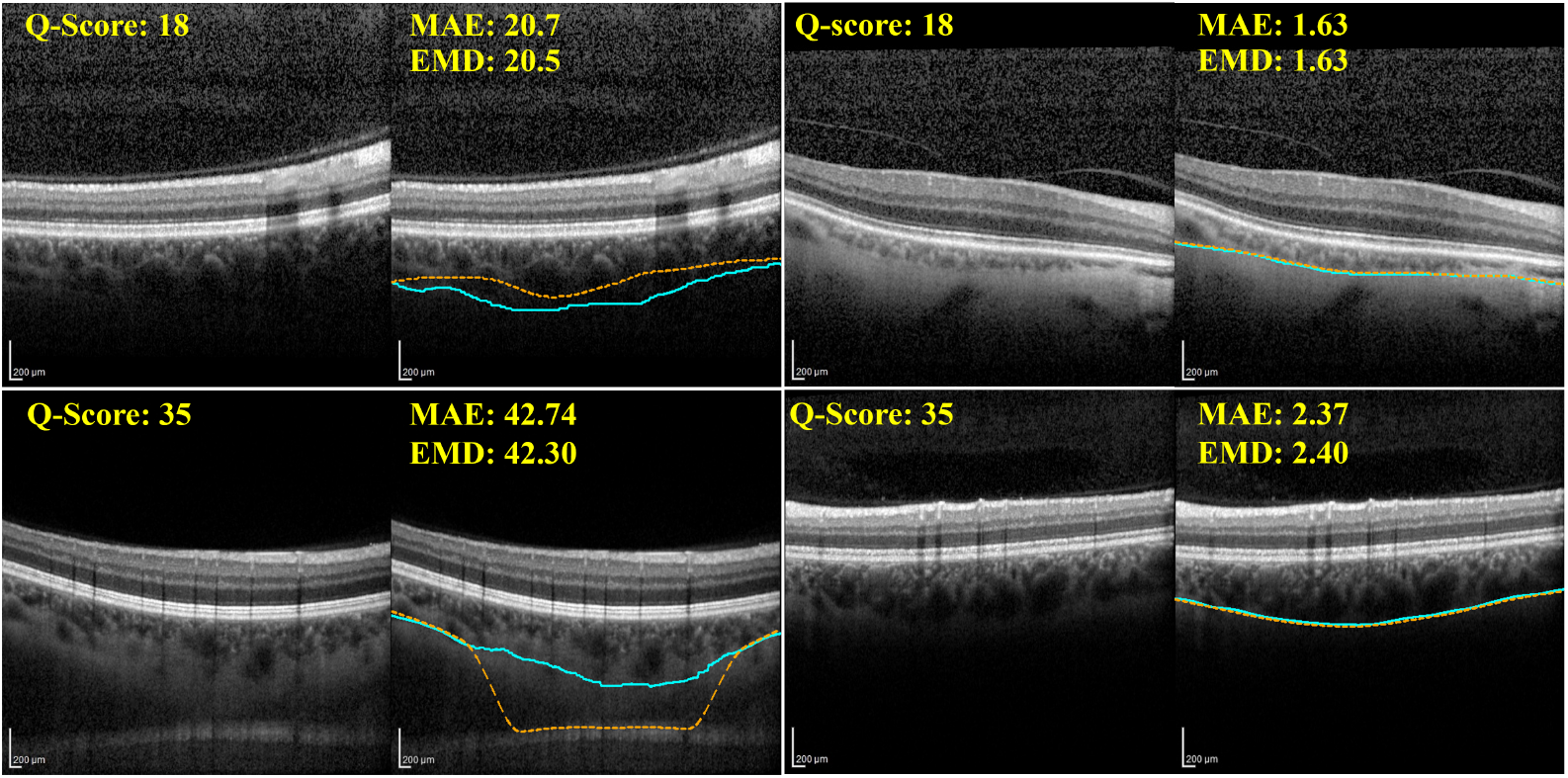
At the COB, higher Q-scores correspond to larger segmentation errors — the opposite of the expected direction. Each row presents one successful and one failed segmentation at the same Q-score level: low (Q-score = 18, top row) and high (Q-score = 35, bottom row). Within each example, the left panel shows the original B-scan and the right panel shows the segmentation overlay. Ground truth: solid cyan. Model prediction: dashed orange. EMD values quantify geometric boundary displacement per overlay. The paradoxical direction reflects anatomical difficulty at the choroid–sclera interface, not image signal strength, as established in the mediation analysis in this section.

The third line of evidence is the depth gradient itself. Segmentation error increases monotonically with anatomical depth — EZ (MAE: 0.87 px; EMD: 1.55 px), BM (MAE: 4.07 px; EMD: 4.51 px), COB (MAE: 6.39 px; EMD: 6.84 px) — while Q-score correlations show no corresponding increase, remaining within |*r*| ≤ 0.117 throughout. Two independent metric families converging on identical boundary ordering confirm this reflects a genuine property of boundary localizability, not an artifact of any particular error formulation. Label noise cannot account for this pattern: ground truth was delineated primarily by a single expert, so any residual annotation variability operates without systematic relationship to depth and cannot explain a monotonic gradient spanning three anatomically distinct boundaries. The supported explanation is anatomical complexity. Boundaries deeper in the tissue lose the local contrast cues that both human graders and learned convolutional features rely on to localize consistently. This difficulty is orthogonal to global signal strength. The Q-score cannot measure it because it does not exist at the level of the image — it exists at the level of the anatomy.

Our findings contrast with those of Muthusivarajan et al., who reported positive correlations between MRI image quality metrics and segmentation Dice scores [31]. We propose this discrepancy reflects fundamental differences in modality, structure, and task. MRI quality metrics are multi-dimensional, capturing resolution, contrast-to-noise ratio, motion artifacts, and field homogeneity. The Q-score, by contrast, is a single scalar — and that distinction matters most at the scale of the segmentation target. MRI segmentation targets large organ volumes where global signal quality is directly relevant; retinal boundary segmentation operates at single-pixel interfaces where local edge contrast matters far more than global brightness. Region segmentation benefits from global quality improvements. Boundary segmentation does not — it depends on local edge properties a global score cannot represent. This distinction is further supported by natural image recognition, where model performance can be largely quality-agnostic yet fails selectively on high-quality images with specific structural properties [32].

These results carry direct implications for the governance of AI-assisted OCT analysis. The Q-score ≥ 15 threshold currently used as a minimum criterion for automated analysis was designed to reflect human-interpretable image quality, not to certify the reliability of deep learning outputs. A scan may be clinically readable yet yield unreliable automated segmentation; conversely, a moderate-quality scan may support accurate boundary localization. Relying on a signal-strength proxy as a quality gate therefore constitutes a silent failure risk. Task-specific acquisition quality criteria are needed — metrics calibrated to predict accurate boundary localization for a given model architecture and a given boundary rather than to reflect global signal interpretability. When the patient is still present, an objective segmentation-likelihood metric could guide operators to rescan immediately — avoiding the clinical cost of recalling the patient for a repeat acquisition. Recent OCT quality work has begun to move in this direction, proposing task-aware metrics that flag scans likely to fail downstream analysis [15]; the acquisition-time application of such metrics remains an open direction that this work motivates. The same principle scales to the population level: distribution shift between training and deployment populations is a widely acknowledged driver of AI model degradation [33], and a task-calibrated acquisition baseline could serve as a drift detection gate without requiring model retraining. In any workflow where AI is involved in informing clinical measurements, a quality gate that does not predict model performance provides no protection against systematically degraded outputs entering the analysis pipeline.

In summary, these findings establish that device-reported signal strength is poorly related to deep learning boundary segmentation performance across all three anatomical boundaries examined. The failure mode is structural — the Q-score measures a global image property while segmentation difficulty is determined by local contrast at specific anatomical interfaces, a mismatch no threshold adjustment can resolve. Clinical deployment of AI-assisted OCT analysis therefore requires a fundamental shift from signal-based to task-calibrated acquisition quality standards.

Limitations of this study include the following. The dataset comprised healthy eyes from a single site, and generalizability to pathological retinas and other DL architectures remains to be established; however, negligible effect sizes across all metric families make a reversal unlikely, as structural disruption would only add variance that a scalar signal-strength index is less equipped to capture. Residual annotator uncertainty at the COB may independently inflate error at that boundary, though it cannot account for the monotonic depth gradient spanning all three boundaries. Future work should examine Q-score–performance relationships in disease cohorts and evaluate whether task-specific, model-calibrated acquisition criteria can replace signal-strength gating in any clinical or research pipeline where AI informs boundary measurements.

## Data Availability Statement

The dataset underlying the results presented in this paper is not publicly available. Requests for data access will be considered on a case-by-case basis and may be directed to the corresponding author.

